# HIV-related knowledge and sexual decision-making indicators among Kenyan boys aged 11–14 years who had heard of HIV: A secondary analysis of the 2018 Kenya Population-based HIV Impact Assessment

**DOI:** 10.64898/2026.10.22.26366194

**Authors:** Erick Ndenga, Cyrus G Kagecha, Nissily Mushani, Pamela M. Murnane, Collins Ouma

## Abstract

Adolescents remain a priority population in HIV prevention efforts in sub-Saharan Africa. Yet, evidence on the relationship between HIV knowledge and sexual decision-making indicators among boys in early adolescence in Kenya is limited. This study examined the association between HIV-related knowledge and intention to have sex before marriage, perceived ability to refuse unwanted sex, and communication with a parent or guardian about sex among Kenyan boys aged 11–14 years who reported knowing what sex was and having heard of HIV.

We conducted a secondary cross-sectional analysis of data from the Kenya Population-based HIV Impact Assessment 2018–2019 (KENPHIA), a nationally representative household survey. The analysis was restricted to boys aged 11–14 years who participated in the child survey, completed the interview, had a valid survey weight, reported knowing what sex is, and had heard of HIV, resulting in a final analytic sample of 495 participants from 1,024 boys who completed the child questionnaire and had valid survey weights. HIV-related knowledge was assessed using a study-specific index derived from five knowledge items and grouped as 0–2, 3–4, or 5 correct responses. The outcomes were intention to have sex before marriage (n = 408), perceived ability to refuse unwanted sex (n = 479), and communication with a parent or guardian about sex (n = 495). Survey-weighted Poisson regression models with a log link and jackknife variance estimation were used to estimate adjusted prevalence ratios (aPRs) and 95% confidence intervals (CIs). Models were adjusted for age, education level, and residence. Sensitivity analyses were conducted by modeling the HIV knowledge score as a continuous variable.

After adjustment for age, residence, and education, HIV-related knowledge categories were not associated with intention to have sex before marriage. Compared with boys with 0–2 correct responses, those with 3–4 or all five correct responses were more likely to report being able to refuse unwanted sex, although prevalence did not rise further in the five-correct group (aPR = 1.32, 95% CI: 1.11–1.57; aPR = 1.29, 95% CI: 1.06–1.57, respectively). Parent/guardian communication prevalence rose across knowledge categories, but category-specific associations were not statistically significant. In sensitivity analyses, each additional correct response was associated with perceived ability to refuse unwanted sex (aPR = 1.08, 95% CI: 1.02–1.14) and parent/guardian communication (aPR = 1.28, 95% CI: 1.03–1.60), but not with intention to have sex before marriage. However, the association for perceived refusal ability departed from linearity.

Among Kenyan boys aged 11–14 years who reported knowing what sex is and having heard of HIV, greater HIV-related knowledge was associated with a higher prevalence of perceived ability to refuse unwanted sex but not with intention to have sex before marriage. Findings for parent/guardian communication differed between the categorical and continuous specifications of the knowledge score and should be interpreted cautiously. Age-specific HIV prevention approaches that combine knowledge with communication, life-skills, safeguarding, and social support interventions warrant prospective evaluation.

## Introduction

Adolescents remain an important population in the global HIV response, particularly in sub-Saharan Africa (SSA), where they continue to experience gaps in HIV-related knowledge and access to age-appropriate prevention information [1]. Despite progress in HIV prevention, treatment and awareness, adolescent boys in particular continue to show gaps in HIV-related knowledge and to engage in behaviors that raise their risk of infection [1]. In 2024, an estimated 1.57 million adolescents aged 10–19 were living with HIV globally, with approximately 82% living in sub-Saharan Africa[2]. During adolescence, boys aged 11–14 years may be forming expectations about relationships and sexual decision-making while still relying on parents, guardians and schools for information. Evidence drawn mainly from older adolescents [3,4] may therefore miss the knowledge and perceptions relevant to this younger group [5]. Understanding these indicators before making assumptions about later behavior can help identify questions for age-appropriate prevention research [6–8].

Although adolescent girls and young women bear a disproportionate burden of new HIV infections in sub-Saharan Africa, adolescent boys also need age-appropriate prevention [1]. A scoping review [9] identified HIV prevention interventions for boys and young men, but noted that their evidence has often been combined with findings for girls or older males. This leaves a need to examine boys in early adolescence as a distinct group.

Accurate knowledge of HIV transmission, prevention and treatment is widely regarded as a foundation for adolescents’ sexual decision-making. However, knowledge alone may not translate directly into behavior because sexual decision-making is also shaped by self-confidence, family communication, peer norms, and the broader social environment. The expanded Health Belief Model formalizes this distinction: knowledge informs perceived susceptibility and the perceived benefits of prevention, but action also depends on perceived barriers and on self-efficacy, the confidence to carry out a protective behavior [10].

The main mode of HIV transmission for adolescents who were not perinatally infected is unprotected heterosexual sex [11]. High-risk sexual behavior, such as early sexual debut, inconsistent condom use, alcohol and drug use, and sensation-seeking, often reinforced by peer pressure, has been associated with increasing HIV infection risk in this age group [12,13].

Comprehensive HIV knowledge, defined as accurate understanding of transmission routes and prevention methods such as condom use and abstinence, together with rejection of misconceptions such as transmission through mosquito bites or shared food, is a cornerstone of HIV prevention because it enables protective behavior [1,14]. However, knowledge gaps persist among adolescents in SSA and are often linked to limited access to age-appropriate sexual education, stigma, and cultural beliefs [15]. Closing these gaps is therefore relevant to the UNAIDS goal of ending AIDS as a public health threat by 2030 [1].

Few studies from sub-Saharan Africa have examined how HIV knowledge relates to sexual risk among boys aged 11–14 years, and work linking early sexual debut to HIV acquisition has given less attention to the intentions and perceptions that develop during early adolescence [16]. Knowledge also does not operate in isolation: boys’ expectations about sex and their perceived ability to refuse unwanted sex may also be shaped by peer norms, access to information and the social environment in which they grow up [5,17]. These influences may be compounded by gender norms that equate risk-taking with masculinity, peer influence, limited access to youth-friendly health care services, and socio-economic disparities [3,18].

The Kenya Population-based HIV Impact Assessment (KENPHIA) 2018 is the most recently completed nationally representative population-level HIV survey with published results available in Kenya. Conducted from June 2018 to February 2019 among individuals aged 0–64 years, it collected comprehensive data on HIV prevalence, incidence, and related knowledge, attitudes, and behaviors, providing a valuable resource for population-level HIV analyses [19]. KENPHIA data reveal gaps in achieving UNAIDS 90-90-90 targets, particularly in the clinical cascade for children above 5 years [20]; however, its behavioral data on young adolescents remain underused. We therefore examined the association between a study-specific HIV-related knowledge score and three indicators among boys aged 11–14 years who reported knowing what sex was and having heard of HIV: intention to have sex before marriage, self-reported ability to refuse unwanted sex, and communication with a parent or guardian about sex.

## Materials and methods

### Study design and data source

We conducted a secondary cross-sectional analysis of data from the Kenya Population-based HIV Impact Assessment (KENPHIA) 2018–2019, a nationally representative household survey designed to estimate HIV prevalence and related health indicators in Kenya. KENPHIA used age-specific questionnaires to collect standardized socio-demographic, HIV-related knowledge, and behavioral data from eligible members of sampled households. This analysis used data from the child questionnaire administered to children aged 11–14 years. A total of 1,024 boys aged 11–14 years completed the individual interview and had valid survey weights. Consistent with the questionnaire skip pattern, eligibility for this analysis was restricted to boys who reported knowing what sex is and having heard of HIV because only these participants were administered the relevant HIV-knowledge questions. Of the 1,024 boys, 541 reported knowing what sex was, of whom 495 reported having heard of HIV and constituted the analytic sample.

Since the questionnaire skip-pattern meant HIV knowledge items were not administered to boys who had not heard of HIV, their scores could not be treated as incorrect. We therefore restricted the analysis to the 495 boys and conducted a sensitivity analysis using the knowledge score as a continuous measure. A further sensitivity analysis was conducted to examine whether having heard of HIV was associated with the study outcomes. Since HIV awareness is related to HIV knowledge, this restriction may have increased the observed knowledge level; the findings apply only to boys who reported knowing what sex was and having heard of HIV and should not be interpreted as estimates for all Kenyan boys aged 11–14 years.

### Variable definition and measurement

Study variables were selected based on the study objectives and the structure of the KENPHIA 2018 child questionnaire. The principal explanatory variable was HIV-related knowledge. The outcome variables were intention to have sex before marriage, perceived ability to refuse unwanted sex, and communication with a parent or guardian about sexual matters. Sociodemographic variables were included to characterize the study population. We selected covariates for multivariable analysis a priori based on their plausible relationships with HIV-related knowledge and the study outcomes. We describe the definitions and coding of all study variables below.

### Exposure: study-specific HIV-related knowledge score

HIV-related knowledge was measured using a study-specific index comprising five questions from the KENPHIA child questionnaire covering HIV prevention, transmission, and treatment. The index was informed by the DHS/UNAIDS comprehensive HIV-knowledge indicators [21]. It was not intended to replace the conventional indicators because it included treatment-related items and did not include all questions concerning common misconceptions.

HIV knowledge was assessed using five items covering prevention, transmission, and treatment: (i) “Can a person reduce their chance of getting HIV by not having sex?”; (ii) “Can a person reduce their chance of getting HIV by using condoms when having sex?”; (iii) “Can a healthy-looking person have HIV or AIDS?”; (iv) “Are there medicines that people with HIV or AIDS can take to help them live longer?”; and (v) “Are there medicines that people with HIV or AIDS can take to make them less likely to spread the virus?” Response options were “Yes”, “No”, “Don’t know” and “Refused”, except for the condom-use item, which offered only “Yes” and “No”. The items were treated as an index of knowledge across related but distinct domains and were not assumed to measure a single latent construct. Correct responses were coded 1 and incorrect or “don’t know” responses were coded 0; refusals were treated as missing. The items were summed to produce a score ranging from 0 to 5, with higher scores indicating more correct HIV-related knowledge. We classified “Don’t know” as incorrect because it indicated an absence of the knowledge assessed by the items.

For ease of presentation, the primary analysis categorized the HIV knowledge scores into 0–2, 3– 4, and 5 correct responses. We defined these categories a priori to facilitate interpretation and ensure adequate numbers of participants in each category. Because these categories were not based on validated clinical thresholds, the HIV knowledge score was also analyzed as a continuous variable in sensitivity analysis to assess the robustness of the findings. Additionally, we assessed the internal consistency of the HIV-related knowledge items using the Kuder–Richardson 20 (KR-20), which was 0.478. Item-rest correlations ranged from 0.188 to 0.442, and removing individual items did not materially improve the coefficient.

Because a continuous specification assumes that each one-point increase has the same association with the outcome across the full 0–5 range, the linearity assumption was assessed by additionally modeling individual knowledge-score values as categorical variables, with score 0 as the reference category. We used a formal joint test to assess whether the score-specific coefficients were consistent with a linear dose-response relationship. Where evidence of non-linearity was observed, the categorical specification was considered more appropriate than imposing a constant one-point effect.

HIV-related knowledge was examined as the principal explanatory variable because greater knowledge of HIV prevention and transmission may be associated with safer sexual intentions, greater confidence in refusing unwanted sex, and increased communication about sexual health among adolescents.

### Outcome variables

Three single-item, self-reported outcomes were examined: (i) intention to have sex before marriage, (ii) perceived ability to refuse unwanted sex, and (iii) communication with a parent or guardian about sex. Intention to have sex before marriage was assessed using the question *“Will you have sex before you get married?*”. Ability to refuse sex was assessed by asking the question, *“If you did not want to have sex with someone, could you tell them that you do not want to have sex with them?”* Parent/guardian communication was assessed by asking the question *“Have you ever talked with a parent or guardian about sex?”.* Each outcome was coded as a binary variable (Yes = 1, No = 0). These measures represent reported intentions, perceptions, and communication rather than observed sexual behavior. Intention to have sex before marriage was not interpreted as a direct measure of HIV-risk behavior because responses may also reflect cultural, religious, and marital expectations. The refusal item assessed a hypothetical, self-reported ability and was not a validated multidimensional measure of sexual self-efficacy.

### Covariates

Covariates were selected a priori based on their plausible associations with both HIV-related knowledge (the principal explanatory variable) and the study outcomes. The multivariable analyses adjusted for age (in years), highest education level attained, and place of residence (urban or rural). These variables were selected because they may influence adolescents’ access to HIV information, educational opportunities, and behavioral intentions. Age was included to account for differences in developmental maturity, education to control for variation in schooling and exposure to health information, and place of residence to account for differences in access to health services and HIV-related information. All covariates were analyzed as categorical variables using the original KENPHIA questionnaire categories.

### Statistical analysis

All analyses were conducted using Stata version 18 (StataCorp LLC, College Station, TX, USA). All analyses accounted for the KENPHIA complex survey design using the KENPHIA final individual interview weight (intwt0). We estimated variance using the 381 KENPHIA jackknife replicate weights (intwt001–intwt381). The survey design was specified in Stata as: svyset [pweight=intwt0], jkrweight (intwt001-intwt381, multiplier (1)) vce(jackknife) mse. Thus, intwt0 was used as the full-sample probability weight, while intwt001 through intwt381 were used to estimate sampling variance using the KENPHIA jackknife replicate-weight procedure. The mean squared error (MSE) option specifies estimation of the variance using the squared deviations of the replicate estimates from the full-sample estimate.

Survey-adjusted generalized linear models were fitted using Stata’s svy: glm command with a Poisson family and log link to estimate prevalence ratios and 95% confidence intervals. Survey-adjusted means, regression analyses of missingness, and cross-tabulations were estimated using svy: mean, svy: regress, and svy: tabulate, respectively.

The primary analytic population consisted of boys aged 11–14 years who had a valid interview and survey weight, reported knowing what sex is, and reported having heard of HIV. This resulted in an analytic sample of 495 participants. For the sensitivity analysis examining HIV awareness itself, the subpopulation was defined among the 541 boys who reported knowing what sex is using Stata’s subpop (if adknsx == 1) specification. This allowed the KENPHIA survey design and replicate weights to be retained when estimating associations within the defined subpopulation.

Descriptive statistics were used to summarize the sociodemographic characteristics of the analytic sample. Categorical variables were presented as unweighted frequencies and survey-weighted proportions with 95% confidence intervals (CIs), while continuous variables were summarized using weighted means, medians, and corresponding 95% CIs where appropriate.

Bivariate analyses were conducted to examine the association between HIV knowledge and each study outcome. Survey-weighted generalized linear models with a Poisson distribution, log link, and design-based standard errors were fitted to estimate prevalence ratios.

The study-specific HIV-related knowledge score was the principal explanatory variable. Multivariable analyses used survey-adjusted modified Poisson regression with a Poisson family and log link to estimate adjusted prevalence ratios (aPRs) and 95% confidence intervals. All models were adjusted for the pre-specified confounders of age, education level, and place of residence, regardless of their statistical significance in bivariate analyses. No formal adjustment for multiple comparisons was applied. The three outcomes represented distinct, pre-specified behavioral and social domains rather than repeated measures of a single primary endpoint. Results were interpreted based on the magnitude and precision of associations, including 95% confidence intervals, rather than statistical significance alone.

As a sensitivity analysis, the HIV-related knowledge score was analyzed as a continuous variable to assess the robustness of the findings. Statistical significance was assessed using a two-sided alpha level of 0.05.

We assessed model stability by examining unweighted outcome event counts across the three HIV-related knowledge categories and by refitting the models after collapsing the three categories into two (scores 0–2 and 3–5).

We conducted design-adjusted joint Wald tests to assess the overall association between the three-level HIV-related knowledge score and each outcome.

We assessed linear trends across the three ordered HIV-related knowledge categories (scores 0–2, 3–4, and 5) by entering the category as an ordinal term in the adjusted survey-weighted Poisson models.

### Missing data

Respondents who reported not knowing what sex is or who had never heard of HIV were not eligible for the HIV-related knowledge assessment under the KENPHIA questionnaire skip pattern and were therefore excluded from the primary analytic sample. The final analytic sample comprised 495 boys aged 11–14 years who reported knowing what sex is and having heard of HIV. Among these participants, HIV knowledge scores were calculable for all respondents, with individual knowledge items scored as correct or incorrect according to the prespecified scoring procedure. Responses indicating incorrect knowledge were scored as incorrect, while refused or otherwise unclassifiable responses were treated as missing, consistent with the study’s coding rules. No refusals or unclassifiable responses occurred on the five knowledge items; therefore, complete scores were calculated for all 495 participants.

Outcome completeness varied across the three outcomes. Data were available for 408 participants (82.4%) for intention to have sex before marriage, 479 (96.8%) for perceived ability to refuse unwanted sex, and all 495 participants (100%) for parent/guardian communication about sex. Missingness in intention to have sex before marriage was not significantly associated with age (p=0.819), education (p=0.100), or HIV-related knowledge category (p=0.228). For perceived ability to refuse unwanted sex, missingness was not significantly associated with age (p=0.329) or education (p=0.527) but was associated with knowledge category (p=0.007). Participants with missing refusal responses had a lower mean HIV knowledge score than those with observed responses (1.75 versus 3.14), suggesting that complete-case estimates for this outcome may be affected by selective missingness. These comparisons should be interpreted cautiously because only 16 participants had missing refusal responses.

### Ethics statement

This study was a secondary analysis of data from the Kenya Population-based HIV Impact Assessment (KENPHIA) 2018. The original survey received ethical approval from the Institutional Review Board (IRB) at the U.S. Centers for Disease Control and Prevention (US CDC), the Columbia University Medical Center IRB, and the Scientific and Ethical Review Unit (SERU) at the Kenya Medical Research Institute (KEMRI) (KEMRI/RES/7/3/1). This secondary analysis was approved by Maseno University Scientific and Ethics Review committee (MUSERC/01470/25.

Maseno University Scientific and Ethics Review committee (MUSERC/01470/25

The secondary dataset used for this analysis was fully de-identified. Access to the KENPHIA/PHIA was obtained through the Population-Based HIV Impact Assessment (PHIA) Data Manager at the International Center for AIDS Care and Treatment Programs (ICAP) at Columbia University, in accordance with the applicable data access, confidentiality and ethical use requirements governing participant-level research data.

## Results

### Study sample

The final analytic sample comprised 495 boys aged 11–14 years who reported knowing what sex is and having heard of HIV. Regression models used complete cases for the outcome and covariates included in each analysis. Consequently, the analytic sample varied by outcome: 375 participants for intention to have sex before marriage, 436 for perceived ability to refuse unwanted sex, and 450 for parent/guardian communication about sex. The participant selection process and outcome-specific exclusions are summarized in Fig 1.

**Fig 1.**
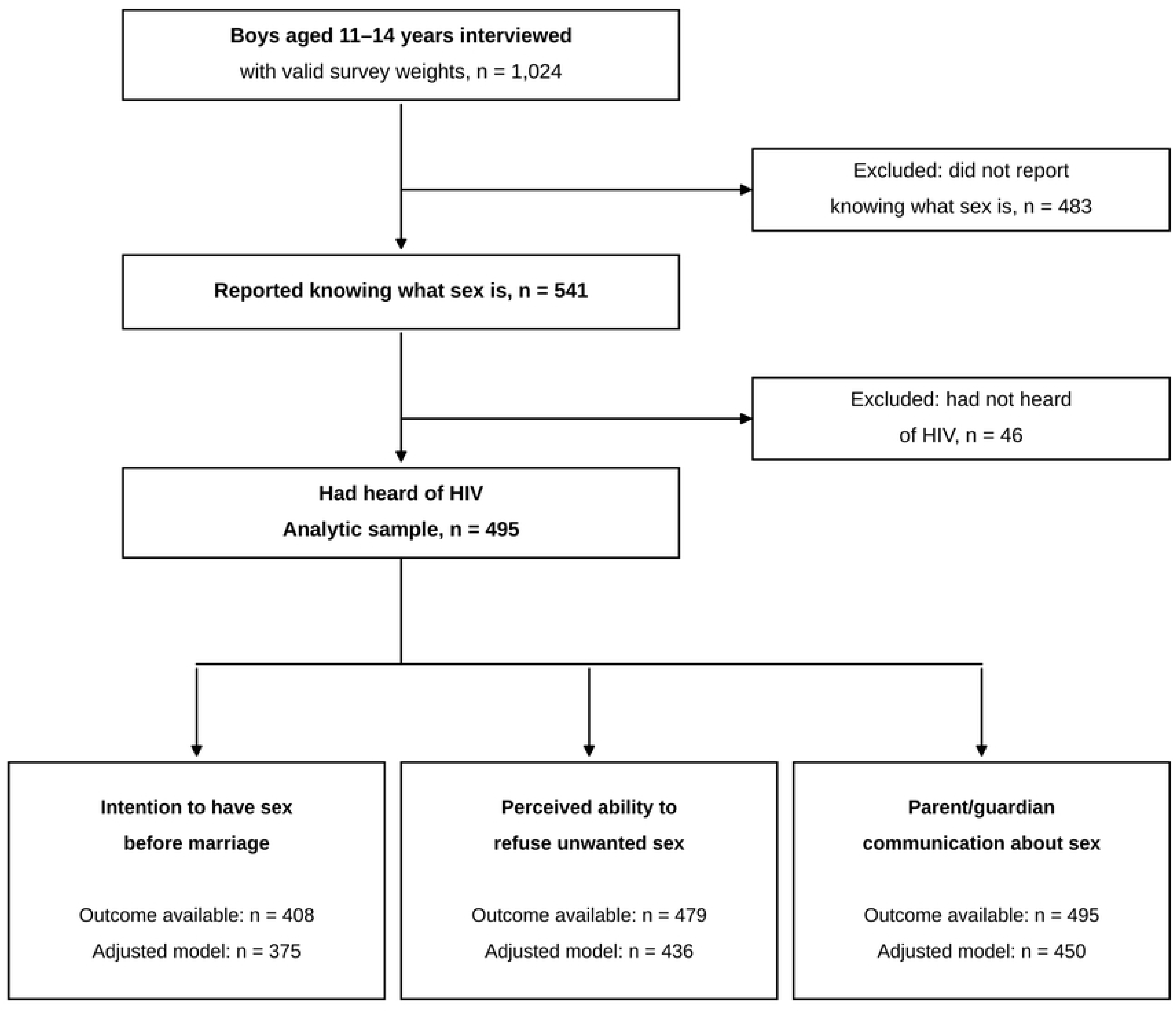
Flow of participants through study eligibility and outcome-specific analyses, KENPHIA 2018–2019.

### Sociodemographic characteristics

The final analytic sample included 495 adolescent boys aged 11–14 years (mean age 12.8 years, 95% CI: 12.7–12.9). Most participants were enrolled in school (94.1%), had primary school as their highest level of education (85.3%), and lived in rural areas (72.1%) (Table 1). Compared with the 529 excluded boys, included boys were older (61.2% vs 34.9% aged 13–14 years) and more often had reached secondary school (8.1% vs 0.2%).

**Table 1.**
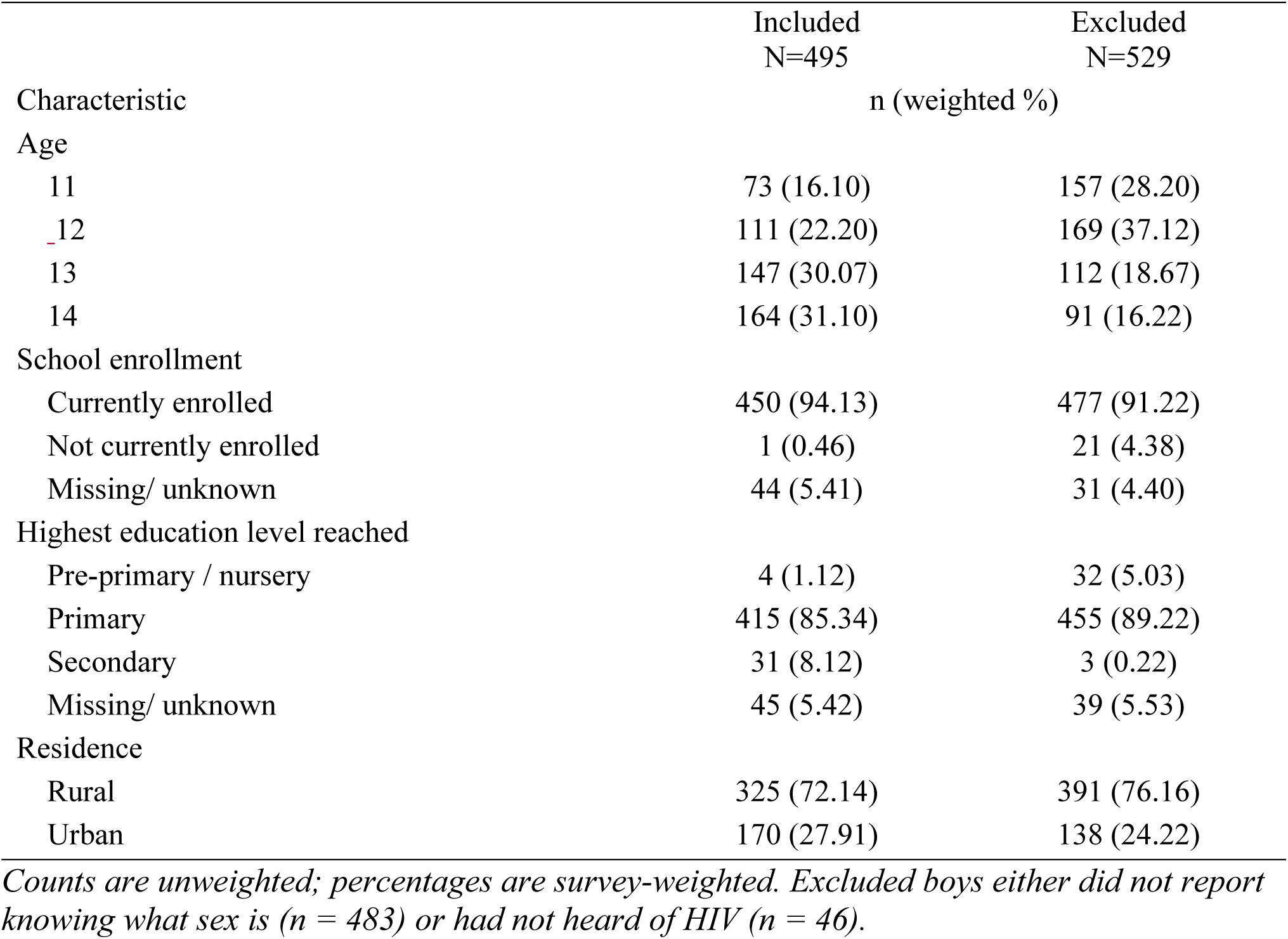
Sociodemographic characteristics of boys aged 11–14 years included in and excluded from the analytic sample, KENPHIA 2018–2019.

### HIV-related knowledge

Correct responses varied across the five knowledge items. The item answered correctly most often was that antiretroviral medicines help people living with HIV live longer (370; weighted 72.8%). Correct responses were also common for recognizing that a healthy-looking person can have HIV (355, 67.6%), that abstinence reduces the risk of HIV infection (322, 65.3%), and that condom use reduces HIV risk (311, 59.9%). The lowest proportion of correct responses was for awareness that antiretroviral therapy can reduce HIV transmission (230, 44.7%). Among the 495 boys in the analytic sample, 135 (30.3%; weighted 95% CI: 25.3–35.3%) scored 0–2, 274 (54.8%; 95% CI: 49.0–60.5%) scored 3–4, and 86 (15.0%; 95% CI: 11.1–18.8%) answered all five items correctly. Thus, the largest proportion scored 3–4 and the weighted mean knowledge score was 3.1 of a possible 5 (SD 1.3; 95% CI: 2.9–3.3).

### Study outcomes

Outcome data were available for 408 participants for intention to have sex before marriage, 479 for ability to refuse unwanted sex, and 495 for communication with a parent or guardian about sexual matters.

Among participants with outcome data, 62/408 (weighted 15.2%) reported intending to have sex before marriage, 359/479 (71.2%) reported that they could refuse unwanted sex, and 78/495 (15.8%) reported having talked with a parent or guardian about sex.

### Association between HIV-related knowledge and study outcomes

Table 2 presents crude and adjusted associations between knowledge category and each outcome.

**Table 2.** Association between HIV-related knowledge score categories and study outcomes among eligible boys aged 11–14 years, KENPHIA 2018–2019.

| Outcome | Knowledge score | n (weighted %) | Crude PR (95% CI) | p-value | Adjusted PR (95% CI) | p-value |
| --- | --- | --- | --- | --- | --- | --- |
| Intention to have sex before marriage |  |  |  |  |  |  |
|  | Score 0–2 (Ref.) | 16 (19.2%) | 1.00 (Ref.) | — | 1.00 (Ref.) | — |
|  | Score 3–4 | 38 (16.1%) | 0.84 (0.42–1.66) | 0.609 | 0.89 (0.44–1.78) | 0.735 |
|  | Score 5 | 8 (10.5%) | 0.54 (0.16–1.84) | 0.327 | 0.61 (0.17–2.11) | 0.429 |
| Perceived ability to refuse unwanted sex |  |  |  |  |  |  |
|  | Score 0–2 (Ref.) | 83 (60.1%) | 1.00 (Ref.) | — | 1.00 (Ref.) | — |
|  | Score 3–4 | 213 (80.2%) | 1.33 (1.13–1.57) | 0.001 | 1.32 (1.11–1.57) | 0.002 |
|  | Score 5 | 63 (75.6%) | 1.26 (1.03–1.53) | 0.024 | 1.29 (1.06–1.57) | 0.010 |
| Communication with parent/guardian about sex |  |  |  |  |  |  |
|  | Score 0–2 (Ref.) | 15 (11.0%) | 1.00 (Ref.) | — | 1.00 (Ref.) | — |
|  | Score 3–4 | 43 (16.1%) | 1.47 (0.73–2.95) | 0.276 | 1.45 (0.70–3.00) | 0.317 |
|  | Score 5 | 20 (22.5%) | 2.06 (0.89–4.75) | 0.091 | 2.19 (0.89–5.40) | 0.088 |
| Notes: Counts are unweighted; percentages are survey-weighted among participants with no missing data for the respective outcome. Crude and adjusted prevalence ratios (PRs) were estimated using survey-weighted Poisson regression with a log link. Adjusted models controlled for age, education and residence. The score 0–2 category was the reference group. Adjusted model sample sizes were 375 for intention to have sex before marriage, 436 for perceived ability to refuse unwanted sex, and 450 for parent/guardian communication. |  |  |  |  |  |  |

In unadjusted analyses, intention to have sex before marriage was not significantly associated with HIV knowledge score category. The survey-weighted prevalence was 19.2% among boys scoring 0–2, 16.1% among those scoring 3–4, and 10.5% among those scoring 5. Compared with boys scoring 0–2, the crude prevalence ratio (PR) was 0.84 (95% CI: 0.42–1.66; p = 0.609) for scores 3–4 and 0.54 (95% CI: 0.16–1.84; p = 0.327) for a score of 5. After adjustment for age, education, and residence, the associations remained non-significant, with adjusted PRs (aPRs) of 0.89 (95% CI: 0.44–1.78; p = 0.735) and 0.61 (95% CI: 0.17–2.11; p = 0.429), respectively.

For perceived ability to refuse unwanted sex, the survey-weighted prevalence was 60.1% among boys scoring 0–2, 80.2% among those scoring 3–4, and 75.6% among those scoring 5. In unadjusted analyses, scores of 3–4 (PR = 1.33, 95% CI: 1.13–1.57; p = 0.001) and 5 (PR = 1.26, 95% CI: 1.03–1.53; p = 0.024) were associated with a higher prevalence of reporting this ability than a score of 0–2. These associations remained statistically significant after adjustment. The aPR was 1.32 (95% CI: 1.11–1.57; p = 0.002) for scores 3–4 and 1.29 (95% CI: 1.06–1.57; p = 0.010) for a score of 5. The findings indicate a higher prevalence of perceived ability to refuse unwanted sex in both higher knowledge-score categories, although the prevalence did not increase consistently across categories.

For communication with a parent or guardian about sexual matters, the survey-weighted prevalence was 11.0% among boys scoring 0–2, 16.1% among those scoring 3–4, and 22.5% among those scoring 5. In unadjusted analyses, neither scores of 3–4 (PR = 1.47, 95% CI: 0.73– 2.95; p = 0.276) nor a score of 5 (PR = 2.06, 95% CI: 0.89–4.75; p = 0.091) was significantly associated with communication. The findings remained non-significant after adjustment, with aPRs of 1.45 (95% CI: 0.70–3.00; p = 0.317) for scores 3–4 and 2.19 (95% CI: 0.89–5.40; p = 0.088) for a score of 5. Although the prevalence estimates were higher in the higher knowledge-score categories, the confidence intervals were wide and included the null value.

Design-adjusted joint Wald tests indicated an overall association between knowledge category and perceived ability to refuse unwanted sex (F(2, 379) = 5.23, p = 0.006), but not intention to have sex before marriage (F(2, 378) = 0.31, p = 0.731) or parent/guardian communication about sex (F(2, 379) = 1.60, p = 0.204). In trend analyses, each one-category increase in knowledge was associated with a 16% higher prevalence of perceived ability to refuse unwanted sex (aPR 1.16; 95% CI: 1.05–1.27; p = 0.002). There was no evidence of a linear trend for intention to have sex before marriage (aPR 0.82; 95% CI: 0.49–1.36; p = 0.431) or parent/guardian communication (aPR 1.48; 95% CI: 0.95–2.31; p = 0.084).

### Sensitivity analyses

To assess the robustness of the primary findings, we modeled HIV knowledge as a continuous score rather than in categories. The models were adjusted for the same covariates used in the primary analysis (age, highest education level and residence) (Table 3).

**Table 3.** Adjusted associations between the continuous HIV-related knowledge score and study outcomes among boys aged 11–14 years, KENPHIA 2018–2019.

| Outcome | Effect | Adjusted PR (95% CI) | P-value |
| --- | --- | --- | --- |
| Intention to have sex before marriage | Per 1-point increase | 0.88 (0.69–1.12) | 0.309 |
| Perceived ability to refuse unwanted sex | Per 1-point increase | 1.08 (1.02–1.14) | 0.005 |
| Parent/guardian communication | Per 1-point increase | 1.28 (1.03–1.60) | 0.028 |

*Adjusted prevalence ratios (PRs) and 95% confidence intervals (CIs) were adjusted for age, highest education level and place of residence (urban/rural). Each PR represents the association between a one-point increase in the HIV knowledge score (range: 0–5) and the respective outcome.*

Consistent with the primary analysis, there was no evidence of an association between HIV knowledge score and intention to have sex before marriage. Each one-point increase in HIV knowledge score was associated with a 12% lower prevalence of reporting this intention (aPR = 0.88, 95% CI: 0.69–1.12; p = 0.309), but the association was not statistically significant.

The positive association between HIV knowledge and the ability to refuse unwanted sex remained evident when HIV knowledge was treated as a continuous variable. Each one-point increase in HIV knowledge score was associated with an 8% higher prevalence of reporting the ability to refuse unwanted sex (aPR = 1.08, 95% CI: 1.02–1.14; p = 0.005), consistent in direction with the categorical analysis. Because this outcome departed from linearity (see below), read the per-point estimate as an average association rather than a constant effect.

In contrast to the primary categorical analysis, the continuous analysis showed a statistically significant association between HIV knowledge and communication with a parent or guardian about sexual matters. For every one-point increase in HIV knowledge score, the prevalence of reporting such communication increased by 28% (aPR = 1.28, 95% CI: 1.03–1.60; p = 0.028). To assess whether a constant one-point association was appropriate, we compared these models with models that treated each score value as a separate category (score 0 as reference) and tested whether the score-specific coefficients were consistent with a linear dose-response relationship.

Design-adjusted Wald tests for departure from linearity found no evidence of departure for intention to have sex before marriage (F(4, 376) = 0.29, p = 0.883) or parent/guardian communication about sex (F(4, 377) = 1.26, p = 0.284), but did find evidence of departure for perceived ability to refuse unwanted sex (F(4, 377) = 3.03, p = 0.018). For this assessment, the knowledge score was modeled categorically, retaining all six values (0–5). Accordingly, the categorical specification was retained for perceived ability to refuse unwanted sex, and continuous-score models were examined as sensitivity analyses. Each one-point increase in knowledge score was associated with a higher prevalence of perceived ability to refuse unwanted sex (aPR = 1.16; 95% CI: 1.05–1.27; p = 0.002), although this estimate should be interpreted cautiously given the evidence of non-linearity. The continuous-score associations were not statistically significant for intention to have sex before marriage (aPR = 0.82; 95% CI: 0.49–1.36; p = 0.431) or parent/guardian communication about sex (aPR = 1.48; 95% CI: 0.95–2.31; p = 0.084).

In an additional sensitivity analysis among the 541 boys who knew what sex was, having heard of HIV was not significantly associated with any of the three outcomes after adjustment for age, education and residence. The adjusted prevalence ratio (aPR) was 0.73 for intention to have sex before marriage (n=413; 95% CI: 0.32–1.64; p=0.440), 1.06 for perceived ability to refuse unwanted sex (n=477; 95% CI: 0.82–1.37; p=0.669), and 0.83 for parent/guardian communication (n=492; 95% CI: 0.32–2.17; p=0.705).

Results from the continuous-score models were consistent with the categorical analyses for intention to have sex before marriage and perceived ability to refuse unwanted sex. In contrast to the categorical analysis, the continuous knowledge score was associated with parent/guardian communication. This difference should be interpreted cautiously because the categorical and continuous models impose different assumptions about the exposure–outcome relationship.

In model-stability checks, the highest-knowledge category (score 5) contained 8 events for intention to have sex before marriage, 63 for perceived ability to refuse unwanted sex, and 20 for parent/guardian communication. After the categories were collapsed into two, the aPR for perceived ability to refuse unwanted sex was 1.32 (95% CI: 1.11–1.56; p = 0.001), close to the three-category estimates. For the other two outcomes, collapsing did not change conclusions about statistical significance, although estimates remained imprecise. One education category contained only four participants, and one jackknife replicate could not be estimated in the intention model; estimates for intention and communication should therefore be interpreted cautiously.

## Discussion

### Main findings

This study used the KENPHIA 2018 data to examine the association between HIV-related knowledge and sexual decision-making indicators among Kenyan adolescent boys aged 11–14 years who had heard of HIV and reported knowing what sex was. Although most participants reported that they were able to refuse unwanted sex, relatively few had discussed sexual matters with a parent or guardian, and a minority reported intending to have sex before marriage.

The principal finding was that boys with 3–4 or 5 correct knowledge responses had a higher prevalence of reporting that they could refuse unwanted sex than boys with 0–2 correct responses. This association remained evident after covariate adjustment and when the categories were collapsed. The continuous-score model pointed in the same direction, although the relationship departed from linearity. Because the study was cross-sectional, the findings do not establish whether greater HIV-related knowledge increases refusal ability or whether both reflect broader exposure to sexuality education, family support, school experiences, or personal confidence.

In contrast, HIV knowledge was not associated with intention to have sex before marriage or communication with a parent or guardian after adjustment for potential confounders. In the continuous-score sensitivity analysis, however, each one-point increase in HIV knowledge score was associated with a higher prevalence of parent/guardian communication about sexual matters. Together, these findings suggest that among these early adolescent boys, HIV knowledge is more strongly associated with psychosocial factors related to sexual decision-making than with behavioral intentions.

### HIV-related knowledge among early adolescent boys

This study found that HIV knowledge among Kenyan adolescent boys aged 11–14 years was moderate, with an average score of 3.1 out of five. However, knowledge was not uniform across the different HIV knowledge domains. Most participants correctly recognized that antiretroviral medicines help people living with HIV live longer and that a healthy-looking person can have HIV. This suggests that many participants understood that treatment can improve survival and that HIV infection cannot necessarily be identified from physical appearance.

In contrast, fewer than half of participants correctly identified that antiretroviral therapy (ART) can reduce HIV transmission, indicating important gaps in understanding newer HIV prevention concepts.

The moderate level of HIV knowledge observed in this study is consistent with findings from a population based survey in Nigeria which found that even though 72% had heard of HIV, only 9.4% met the study’s specific definition of comprehensive HIV Knowledge[22]. Adolescents have also been reported to be more familiar with traditional HIV prevention messages, such as abstinence and condom use, than with more recent information related to HIV treatment and treatment as prevention [23]. This may reflect the greater emphasis that school-and community-based HIV education programs have historically placed on preventing HIV acquisition rather than explaining advances in HIV treatment [24].

The relatively low level of knowledge regarding the role of ART in reducing HIV transmission is particularly important because treatment as prevention has become a central component of HIV control programs. Although this concept is widely promoted within HIV care settings, younger adolescents may have fewer opportunities to receive detailed information on HIV treatment compared with older adolescents or adults. Strengthening HIV education to include age-appropriate information on HIV treatment, viral suppression, and treatment as prevention may therefore improve adolescents’ overall understanding of HIV prevention and possibly improve psychosocial and behavioral-intention outcomes.

### HIV knowledge and the ability to refuse unwanted sex

Higher HIV knowledge was associated, after adjustment, with a greater reported ability to refuse unwanted sex among adolescent boys aged 11–14 years who knew what sex was and had heard of HIV. Compared with boys with 0–2 correct responses, those with 3–4 or 5 correct responses had an approximately 30% higher prevalence of reporting that they could refuse unwanted sex after adjustment for age, education level, and place of residence. The association persisted when categories were collapsed, and the continuous-score model (aPR 1.08 per point) pointed in the same direction, although prevalence did not rise further between the 3–4 and 5 categories. Together, these findings suggest a positive but not strictly dose-dependent relationship between HIV knowledge and perceived refusal ability during early adolescence.

This finding is consistent with previous studies conducted among adolescents in sub-Saharan Africa, which have shown that greater HIV knowledge is associated with improved self-efficacy, stronger decision-making skills, and increased confidence to adopt protective sexual behavior [25,26]. Although many studies have focused on older adolescents, the present findings suggest that similar associations may be observable during early adolescence. This is an important observation because early adolescence represents a critical developmental period during which beliefs, attitudes, and behavioral expectations are formed [27].

Several explanations may account for this association. Greater HIV-related knowledge may occur alongside exposure to sexuality education that also addresses assertiveness and decision-making. Alternatively, adolescents with stronger communication skills, supportive family or school environments, or greater personal confidence may be more likely both to acquire HIV knowledge and to report that they could refuse unwanted sex. These unmeasured factors may partly explain the observed association.

This pattern is consistent with the expanded Health Belief Model, under which individuals are more likely to adopt protective behaviors when they possess both the knowledge and confidence needed to act. The observed association between HIV knowledge and the ability to refuse unwanted sex is compatible with strengthening HIV knowledge before sexual behaviors become established, although the cross-sectional design cannot show that knowledge precedes refusal ability.

From a public health perspective, these findings highlight the value of introducing comprehensive HIV knowledge during early adolescence. Strengthening HIV knowledge before sexual debut may help equip boys with the knowledge and confidence needed to make safer decisions as they transition into later adolescence [9,28]. HIV prevention efforts should combine factual HIV-related knowledge with communication, negotiation, and refusal skills to support healthy adolescent development.

### HIV knowledge, parent/guardian communication, and intention to have sex before marriage

Unlike the ability to refuse unwanted sex, HIV knowledge was not associated with communication about sexual matters with a parent or guardian or with intention to have sex before marriage in the adjusted analyses. However, when HIV knowledge was analyzed as a continuous score, each one-point increase in HIV knowledge was associated with a 28% higher prevalence of parent/guardian communication. This suggests that although categorizing HIV knowledge into broad levels did not detect a statistically significant association, increasing HIV knowledge may still be associated with greater communication between adolescents and their parents or guardians.

Communication between parents and adolescents is widely recognized as an important component of HIV prevention and healthy adolescent development. Previous studies have shown that open discussions about sexuality are associated with improved HIV knowledge, delayed sexual initiation, and safer sexual behaviors among adolescents [28,29]. Despite these benefits, only 15.8% of participants in the present study reported having discussed sexual matters with a parent or guardian. This low level of communication is consistent with findings from many sub-Saharan African countries, where conversations about sexuality within families are often limited by cultural norms, embarrassment, and inadequate parental knowledge or confidence to discuss sexual health with younger adolescents [30–35].

The absence of a statistically significant association in the primary analysis suggests that HIV knowledge alone may not be sufficient to promote parent/guardian communication. Communication within families likely depends on broader social and family factors, including parental attitudes, cultural expectations, religious beliefs, household relationships, and opportunities for discussion. As a result, programs that seek to improve adolescent HIV knowledge may achieve greater impact if they also include interventions that strengthen parents’ skills and confidence to discuss sexual and reproductive health with their children.

Similarly, HIV knowledge was not associated with adolescents’ intention to have sex before marriage. This finding suggests that behavioral intentions during early adolescence may not be determined by HIV knowledge alone, consistent with the Health Belief Model’s emphasis on perceived barriers, benefits and self-efficacy rather than information [10]. Responses to this item may also reflect religious and marital norms rather than anticipated risk.

Behavioral intentions are influenced by a complex interaction of individual, family, peer, school, and community factors. During early adolescence, many boys are still developing their attitudes, beliefs, and expectations regarding relationships and sexuality. Consequently, while HIV knowledge provides important factual information, intentions related to future sexual behavior are also shaped by social norms, parental supervision, peer influences, religious values, and the wider environment [29,33,36]. These findings are consistent with evidence that knowledge is an essential component of HIV prevention but is unlikely to produce meaningful behavioral change when implemented in isolation.

Overall, the findings suggest that HIV knowledge programs targeting early adolescent boys should extend beyond improving knowledge alone. Combining age-appropriate HIV education with interventions that strengthen parent/guardian communication, build supportive family environments, and promote life skills may provide a more comprehensive approach to supporting healthy sexual development during early adolescence.

### Strengths and limitations

This study used data from a large national household survey and accounted for the complex survey design using sampling and replicate weights. It also examined HIV-related knowledge across prevention, transmission, and treatment domains and compared findings using categorical and continuous specifications of the knowledge score.

Several limitations should be considered. First, the cross-sectional design precludes determination of temporality or causality. Reverse causation is particularly plausible for parent/guardian communication, which may be a source as well as a correlate of HIV knowledge. Second, the analysis was restricted to boys who reported knowing what sex was and having heard of HIV. These boys were generally older and more educated than excluded boys; therefore, the findings should not be generalized to all Kenyan boys aged 11–14 years and may be affected by selection bias. Third, the study-specific knowledge index had low internal consistency (KR-20 = 0.48) and its categories were not validated thresholds, and the outcomes were measured using single self-reported items, including a hypothetical measure of refusal ability. Fourth, outcome-specific missing data reduced the available samples and may have introduced bias if missingness was systematic. Fifth, the data were collected in 2018–2019 and may not reflect current knowledge levels or programming. Finally, social-desirability bias, residual confounding, and the examination of multiple outcome–exposure specifications should be considered when interpreting the findings. Despite these limitations, the study provides meaningful evidence concerning associations between HIV-related knowledge and selected sexual decision-making indicators among Kenyan boys 11–14 years old who already knew what sex was and had heard of HIV.

### Implications for policy and practice

The findings of this study have important implications for HIV prevention programs targeting early adolescent boys in Kenya. First, the observed association between higher HIV knowledge and the ability to refuse unwanted sex supports prospective evaluation of interventions combining HIV education with age-appropriate communication and refusal-skills components. Because questions about unwanted sex among children may involve coercion, abuse, or exploitation, programs should incorporate confidential, age-appropriate safeguarding and referral procedures. Upper primary school, when many adolescents are beginning to form attitudes related to sexual health, is a logical setting for such evaluation.

Second, the moderate HIV knowledge observed in this study, together with the low level of knowledge regarding the role of antiretroviral therapy in reducing HIV transmission, highlights the need to strengthen the content of HIV education programs. HIV education should extend beyond traditional prevention messages to include age-appropriate knowledge on HIV treatment, viral suppression, and current HIV prevention strategies. Ensuring that adolescents receive accurate and comprehensive information that incorporates safeguarding approaches may better prepare them to make informed decisions as they progress through adolescence.

Third, although HIV knowledge was associated with greater confidence to refuse unwanted sex, communication about sexual matters with parents or guardians remained low. These findings suggest that school-based HIV education alone may be insufficient to support healthy sexual development. Interventions that actively engage parents and caregivers and strengthen their ability to communicate with adolescents about sexual and reproductive health may complement school and community programs and create a more supportive environment for HIV prevention.

Finally, the findings reinforce the importance of delivering comprehensive HIV prevention interventions that combine knowledge with life-skills development, communication skills, and supportive family and community environments. Such interactive approaches that strengthen decision-making, communication, and resistance-to-pressure skills may be more effective than information-based interventions alone in promoting healthy behaviors among early adolescent boys.

## Conclusion

Among Kenyan adolescent boys aged 11–14 years who had heard of HIV and knew what sex was, higher HIV knowledge was associated, after adjustment for age, education and residence, with a higher reported ability to refuse unwanted sex. However, HIV knowledge was not associated with intention to have sex before marriage. Findings for parent/guardian communication differed between the categorical and continuous specifications of the knowledge score and should therefore be interpreted cautiously.

These results indicate that HIV-related knowledge may be associated differently with specific psychosocial and behavioral-intention outcomes during early adolescence. Prospective studies using validated knowledge and self-efficacy measures are needed to establish temporality and evaluate whether age-appropriate programs combining HIV education, communication, life skills, and safeguarding support improve subsequent outcomes.

## Data Availability

No data were generated by this study. The following existing data source was used: the Kenya Population-based HIV Impact Assessment (KENPHIA) 2018–2019 dataset, available through the PHIA Data Manager hosted by ICAP at Columbia University. Access requires electronic registration and agreement to the data-use terms. Code used can be made available on request.

## Acknowledgments

We thank the National AIDS and STI Control Programme (NASCOP), the KENPHIA survey teams and participants, and ICAP at Columbia University for providing access to the KENPHIA data.

## Notes

### Competing Interest Statement

The authors have declared no competing interest.

### Author Declarations

This study was a secondary analysis of data from the Kenya Population-based HIV Impact Assessment (KENPHIA) 2018. The original survey received ethical approval from the Institutional Review Board (IRB) at the U.S. Centers for Disease Control and Prevention (US CDC), the Columbia University Medical Center IRB, and the Scientific and Ethical Review Unit (SERU) at the Kenya Medical Research Institute (KEMRI) (KEMRI/RES/7/3/1). This secondary analysis was approved by Maseno University Scientific and Ethics Review committee (MUSERC/01470/25. Maseno University Scientific and Ethics Review committee (MUSERC/01470/25

